# Taiwan on track to end third COVID-19 community outbreak

**DOI:** 10.1101/2021.06.20.21259178

**Authors:** Torbjörn E. M. Nordling, Yu-Heng Wu

## Abstract

Since the start of the COVID-19 pandemic on December 31st, 2019, with the World Health Organization being notified of pneumonia of unknown cause in Wuhan (China), Taiwan has successfully ended two COVID-19 community outbreaks. For 19 days, the third community outbreak has now been successfully suppressed, putting Taiwan on path to end it too around Aug. 16th based on our forecast using an exponential model. Since May 28th the 7-day average of reported confirmed infected, which peaked at 593, has been falling to 204 on June 16th and the 7-day average of reported suspected and excluded cases increased to above 25 000. Resulting in a decrease in the ratio of the 7-day average of local & unknown confirmed to suspected cases–the identified control variable–to less than one third of its peak value. The later is a hallmark of working contact tracing, which together with testing and isolation of infected are the keys to ending the community outbreak.

## Introduction

The first outbreak started a little before February 16th and ended April 11th, 2020 (54 days), see Figure 1a. The second outbreak started a little before January 12th and ended February 9th, 2021 (27 days). The third outbreak started a little before April 20th and is still ongoing (56 days), but shows clear signs of having peaked on May 28th. The record of 721 confirmed locally or unknown acquired infections in a day was reported on May 22nd. Between the first and second outbreak Taiwan saw a period with 273 days without any reported local infection, excluding an isolated case of unknown source reported August 2nd and a local case reported December 22nd. Between the second and third outbreak Taiwan had 69 days without any reported local infection.

**Figure 1:**
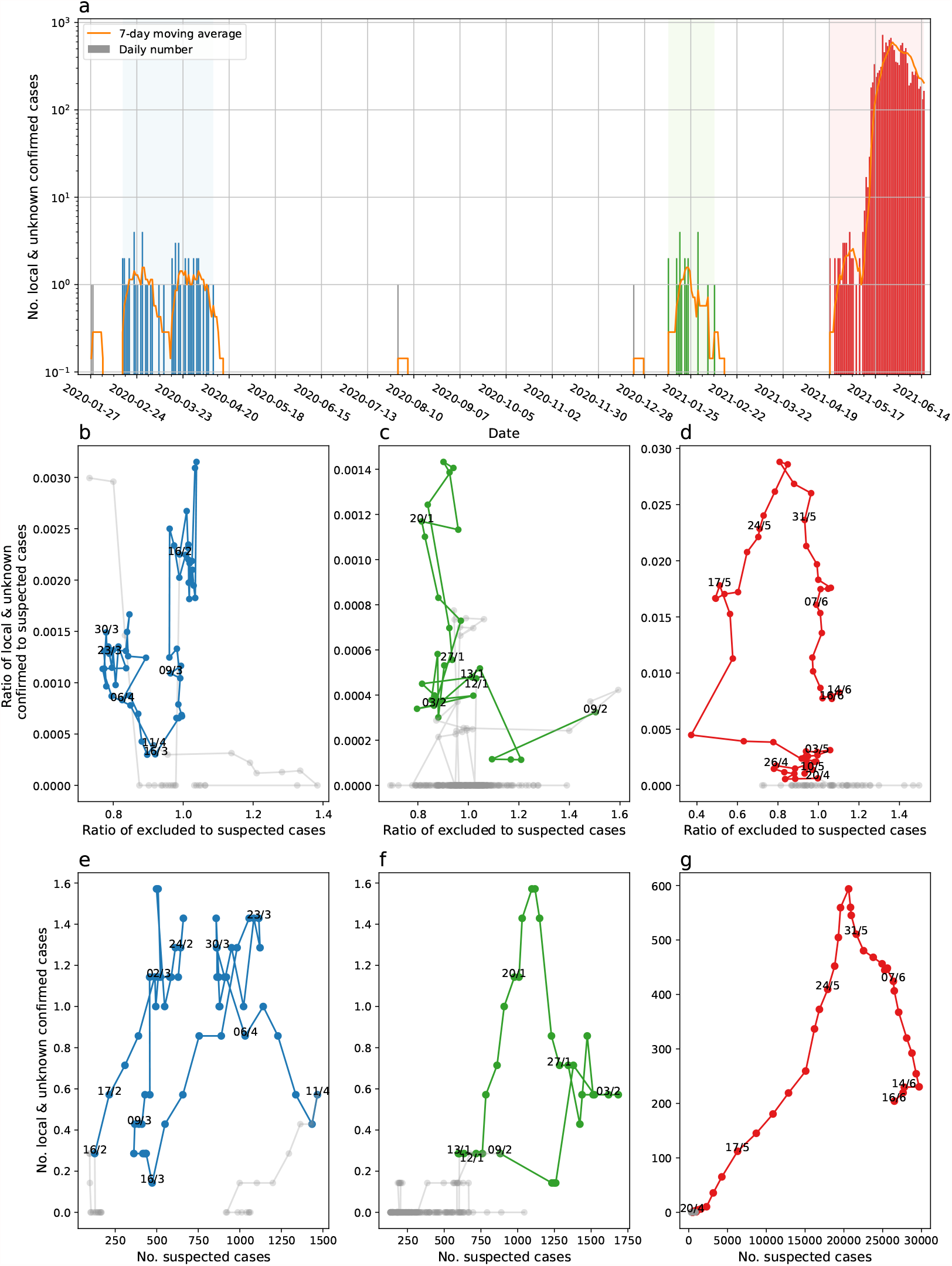
Reported number of local and unknown infected and 7-day moving average in Taiwan from the first local case 2020-01-28 until 2021-06-16 (a). The three community outbreaks are marked in blue (1st), green (2nd), and red (3rd). The cases between are not connected. The ratio of local and unknown confirmed to suspected cases versus ratio of excluded to suspected cases reported for the three outbreaks (b-d). Number of local and unknown confirmed cases versus number of suspected cases reported for the three outbreaks (e-g). The 7-day moving average of each quantity is used in b-g.

## Results and Discussion

From June 2020 until the current outbreak, life in Taiwan was pre-pandemic, except for the mandatory mask and temperature checks in the public transportation introduced on March 31st^1^ and at large events. Large events with tens of thousands of visitors have been held, such as the Mayday New Year’s Eve pop concert at Taoyuan International Baseball Stadium on December 31st^2^. Both the 2nd and 3rd outbreak involved the alpha variant (B.1.1.7), which Hsu et al.^3^ estimated to have a 1.44-fold higher infection probability and 57% higher basic reproduction number based on household transmissions during the 1st and 2nd outbreak, in agreement with previous estimates of 43-90%^4^. Evidently, the measures in place to reduce transmissibility and prevent a large scale outbreak were not sufficient, so Taiwan should have had a surge in cases if community transmission existed during the two periods without local infections. In addition, the National Health Insurance Administration (NHIA) has proactively been seeking out patients with severe respiratory symptoms in its database and allowing all hospitals, clinics, and pharmacies to see the patients’ travel history obtained from the National Immigration Agency from February 18th, 2020 onwards^5^. The current outbreak has been traced to a cluster of infected China Airline pilots and the Novotel at Taoyuan International Airport violating the quarantine rules by housing quarantined flight crews and local guests in the same building^6,7^. This initial failure of the mandatory quarantine at the boarder and contact tracing during the current outbreak combined with the success of contact tracing during the 2nd outbreak and the transmissibility reduction measures remaining the same, implies that Taiwan’s success is due to border control and tracing of contacts upon suspicion. Thus we can finally with confidence say that Taiwan has ended two community outbreaks thanks to contact tracing, testing, and isolation.

The Taiwanese COVID-19 control strategy implemented by the Central Epidemic Command Center (CECC), activated on January 20th, 2020^5^, is based on six main pilars: (a) border control with quarantine upon arrival, (b) self-health monitoring when having visited a place with known cases, (c) testing when showing symptoms and seeking medical care, (d) mandatory supervised quarantine of confirmed infected and individuals at high-risk of having been infected, (e) contact tracing, and (f) a four-level system of measures to suppress community spreading. These are devised to control the effective reproduction number, i.e. the expected number of people an infected individual will transmit the disease to while infectious. The reproduction number can be seen as the transmission risk per contact (transmissibility), times the number of contacts per day, times the number of days the person is infectious. From June 7th until May 11th no restrictions on the size of gatherings, i.e. curbing of the number of contacts, existed^8,7^. The current level 3 measures in place since May 19th, consist of mandatory wearing of masks at all times outside private spaces and social distancing, which reduce the transmissibility; indoor gatherings limited to five people and closure of certain businesses, partly including schools and preschools, which reduce the number of contacts; and mandatory COVID-19 testing in areas where community transmission has taken place, which reduces the number of days an infected person can transmit the disease before being quarantined^9,10^. As demonstrated by numerous countries, such as the United Kingdom and United States, community suppression measures, such as lockdowns, alone do not end a community outbreak. As soon as these non-pharmacological interventions (NPIs) are lifted the reproduction number will increase again, so why do we claim that Taiwan is on track to ending this community outbreak?

In short, the ratio of the 7-day moving average of confirmed infected to suspected has already fallen to less than one third of the peak value of 0.029 on May 28th, while the number of daily suspected has increased to above 25 000, and the ratio of the 7-day moving average of excluded to suspected is slightly above one, which is needed to resolve the backlog from when it was down at 0.37. In other words, the contact tracing, testing, and isolation of infected has, after initial failure and challenges, successfully been scaled up to the required volume of around 25 000 suspected per day and the number of confirmed infected is decreasing. The same pattern has been repeated during all three outbreaks: first the number of confirmed infected increase rapidly and the contact tracing, testing, and exclusion of cases lags, then focus on contact tracing brings up the number of suspected and investment into expanding testing clears the backlog, see Figure 1b-g. Thus the key to ending an outbreak is to focus on contact tracing to bring up the number of suspected and increase testing and isolation/care capacity. The focus on contact tracing and bringing up the number of suspected is key because one need to prevent every infected person from infecting a new person to end the outbreak. It makes the ratio of confirmed infected to suspected the essential metric to follow. The Lancet Commission Task Force on public health measures to suppress the pandemic also report on the effectiveness of contact tracing and quarantine measures, albeit not on its use to end outbreaks^11^. Since May 19th, Taiwan has implemented an SMS text message contact tracing system based on each person upon entry in each business scanning a place specific QR code that generates a unique SMS sent to 1922–the Taiwan Center for Disease Control hotline^12^, which helps scale the contact tracing. This data tells us that the contact tracing, testing, and isolation capacity has been scaled to the required level. Therefore, it is not a question of if the outbreak can be ended but a question of when it will be ended. It is a matter of keeping up the work until this outbreak also is brought to an end. To keep up the work requires will–will from everyone to contribute to the contact tracing, self-health monitoring, testing, and caring of each other. To have will, one need to believe that it can be done, therefore it is vital to get this message to everyone: Together we can and will also end this outbreak.

The two extensions of the level 3 measures have made some ask if vaccination is necessary to end this outbreak? Of the only three countries, with a population over 1 million, that have vaccinated over half their population, both Bahrain and Mongolia in the past week reported over 5 000 confirmed infected, while Israel reported a mere 117 infected^13^. Actually, both Bahrain and Mongolia have set new records in the weekly number of confirmed infected within the previous three weeks. Israel only recently, on June 1st, lifted most restrictions, including limitations on the number of people at gatherings and proof of vaccination to enter e.g. gyms, theatres, hotels, and synagogues, and have kept border controls^14^. In a recent simulation study, Moghadas et al.^15^ predict that if all unvaccinated individuals in the United States revert to pre-pandemic life on July 4th, then it would result in the worst daily incidence of COVID-19 cases despite assuming close to 70% being vaccinated, but if only the vaccinate individuals revert, then it result in a minimal increase even if they revert two weeks after the first dose. Based on the recent estimate of the basic reproduction number without NPIs in Taiwan during the second outbreak at 6.7^3^, 85% of the Taiwanese population would need to be vaccinated to reach herd immunity. Considering the difficulties the Taiwanese government has faced in purchasing COVID-19 vaccines, it is unlikely that Taiwan could implement a population scale vaccination program and reach herd immunity any faster than other nations. A population wide vaccination campaign is likely to take a year before it could end this community outbreak, while the proven method of contact tracing, testing, and isolation is likely to end it around Aug. 16th. We forecast this end date based on fitting an exponential model to the daily ratios of the 7-day moving average of confirmed infected to suspected from the peak of 0.029 on May 28th to 0.0077 on June 16th. The calculated non-simultaneous prediction bounds on a new observation with 95% confidence puts the earliest end date on July 2nd and the latest indefinitely into the future, see Figure 2. The predicted half-time is 9.9 days (95% confidence interval: 9.1-10.8). We therefore encourage all Taiwanese to be hopeful and remain vigilant; the end around Aug. 16th is only two months away.

**Figure 2:**
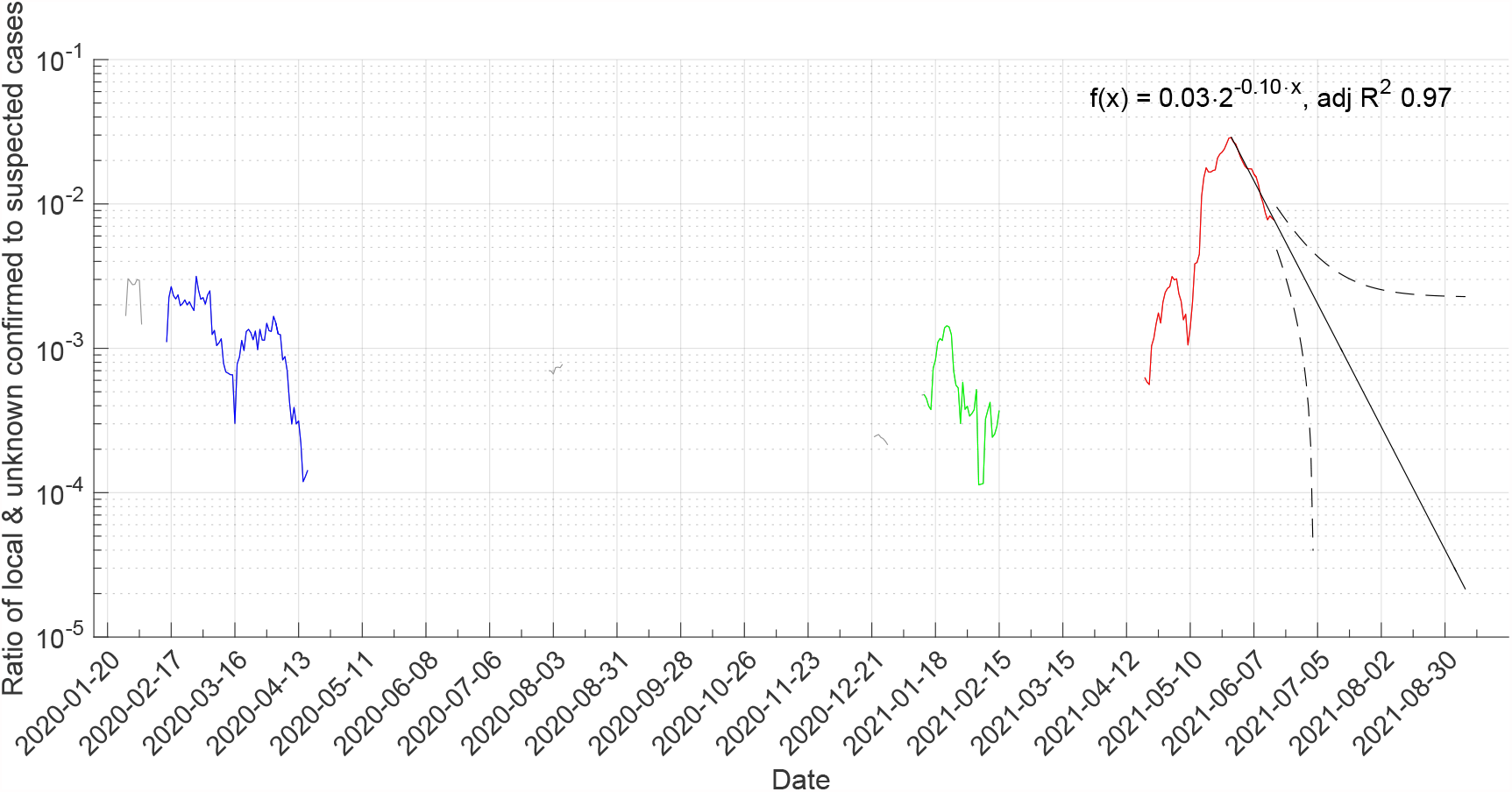
Evolution of the ratio of the 7-day moving mean of local & unknown confirmed to the 7-day moving mean of suspected cases and our forecast of the end of the third outbreak. The three community outbreaks are marked in blue (1st), green (2nd), and red (3rd). The cases between in grey are not connected.

## Conclusion

As soon as this outbreak is ended, we hope the world would take notice and learn from the Taiwanese strategy, so unnecessary suffering and deaths can be avoided. Like Wang et al.^5^, we think “Taiwan is an example of how a society can respond quickly to a crisis and protect the interests of its citizens.”

## Methods

To limit the influence of reporting differences between the week days and fluctuations in when cases are reported to the Taiwan CDC, we use the 7-day moving mean calculated over the past seven days. It is worth noting that our moving mean cover the mean serial interval, which was estimated to 6.2 days among the 42 infector-infectee pairs during the 1st outbreak^18^.

To forecast when the current third community outbreak will end, we assumed that the decrease in the ratio of local & unknown confirmed to suspected cases can be described as an exponential growth process. More precisely, we assumed the model

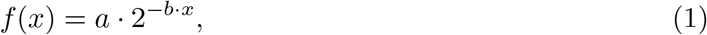

where *a* is the initial value and the reciprocal of *b* is the half-time. We fitted this model to the ratio of the 7-day moving mean of local & unknown confirmed to the 7-day moving mean of suspected cases from the peak on 2021-05-28 to 2021-06-16, i.e. 19 time points, using the Curve Fitting Toolbox in Matlab R2015b by MathWorks using the ”NonlinearLeastSquares” estimator. The root mean squared error of the model fitted to the data points was 0.0011 and adjusted R-squared 0.97, indicating a good fit that explains the data well. The estimated parameters are *a* 0.029 (95% confidence bounds 0.028 - 0.030) and *b* 0.10 (95% confidence bounds 0.09 - 0.11). The later corresponds to a halving of the ratio every 9.9 days (95% confidence bounds 9.1 - 10.8 days). The previous two outbreaks have ended after the ratio has been brought down to 1.2· 10^*−*4^, see Figure 2, so we assume that the same is required to end this outbreak. Note that the length of the window of our moving mean and the baseline number of suspected affects this ratio. When only one confirmed infected remains, the nominator will be 0.14, i.e. 1/7, and the denominator then need to be around 1200, i.e. on average 1200 suspected each day for the last confirmed infected. The mean 7-day moving mean number of suspected between the 1st and 2nd outbreak was 324 and between the 2nd and 3rd outbreak it was 684. By extrapolation we forecast that this ratio would be reached on 2021-08-16 after required six halvings. This forecast depends on the contact tracing, testing, and isolation being equally effective as during the past three weeks and no new local clusters emerging from people infected abroad. The calculated non-simultaneous prediction bounds on a new observation with 95% confidence, drawn as black dashed lines in Figure 2, puts the earliest end date on July 2nd and the latest indefinitely into the future. The later in agreement with a failure to execute the contact tracing, testing, and isolation or new local clusters emerging from people infected abroad.

## Data Availability

All COVID-19 data were collected from the Taiwan Centers for Disease Control (Taiwan CDC), which provided an online platform for downloading datasets, including respiratory syndrome coronavirus-2 (SARS-CoV-2) infection data.
Both data downloaded from the online platform and extracted from their News Bulletins were used.

https://data.cdc.gov.tw/en/dataset/daily-cases-suspected-sars-cov-2-infection_tested

https://www.cdc.gov.tw/Bulletin/List/MmgtpeidAR5Ooai4-fgHzQ

## Acknowledgements

The authors acknowledges valuable discussions, help with presentation, and comments from Dr. Akram Ashyani, Dipesh Dhayfule, and Jose Chang.

## Contribution statement

TEMN did conceptualisation, funding acquisition, methodology, project administration, resources, supervision, and writing of the original draft. YHW did investigation, data curation, formal analysis, and software development. Both did data validation, visualisation, and review & editing of the final manuscript.

## Data

All COVID-19 data were collected from the Taiwan Centers for Disease Control (Taiwan CDC), which provided an online platform for downloading datasets, including respiratory syndrome coronavirus-2 (SARS-CoV-2) infection data^16^. Both data downloaded from the online platform and manually extracted from their News Bulletins^17^ were used. All data used is for the readers convenience also included in the Appendix.

## Appendix

Here we provide details on the data collection, cleaning, and analysis to enable verification and reproduction of the results.

## Data collection

All data is publicly available and was collected from the Taiwan Centers for Disease Control (Taiwan CDC). We collected the cumulative number of confirmed infected with SARS-CoV-2 with a know local source and unknown source, as well as the cumulative reported suspected and excluded cases, from the Taiwan CDC News Bulletins, available in Chinese at https://www.cdc.gov.tw/Bulletin/List/MmgtpeidAR5Ooai4-fgHzQ (last visited 2021-06-17). Unfortunately, these news bulletins have not been issued every day, nor at the same time of the day through out the pandemic, which means that the daily increase derived by subtracting the value in the previous report is influenced by the varying time between these reports. The way of counting the number of suspected and excluded was changed on 2020-03-06, which contain a large correction to these cumulative numbers. We therefore also downloaded the ”Daily Number of Cases Suspected SARS -CoV-2 Infection Tested” dataset from the Taiwan CDC Open Data portal https://data.cdc.gov.tw/en/dataset/daily-cases-suspected-sars-cov-2-infection_tested (last visited 2021-06-17). This dataset contains the daily number of cases suspected to have SARS-CoV-2 from three sources: (i) notifications of infectious diseases, (ii) home quarantine and inspection, and (iii) expanded monitoring, as well as the total daily number.

## Data cleaning and preprocessing

We first removed the first CDC News Bulletin for the days when more than one was issued, thus keeping the later report with the larger cumulative numbers. The we corrected for the number of days in between the report for all cases with more than one day in between by evenly distributing the change in cumulative number over the days in between and ensuring that the cumulative sum equals the reported one. Due to the change in the way of counting the number of suspected and excluded on 2020-03-06, the number of suspected cases is unreasonably low before it and a jump correcting for the change occur on that day. These artefacts are clearly seen in Figure 3 as a dip in the number of suspected before and a value slightly above 10 000 on this day. These artefacts clearly influence the 7-day moving mean of the number of suspected that we need for getting a stable estimate of the ratio of confirmed infected to suspected cases. The total daily number of suspected cases in the CDC Open Data portals dataset does not contain this error, so we decided to use it instead.

**Figure 3:**
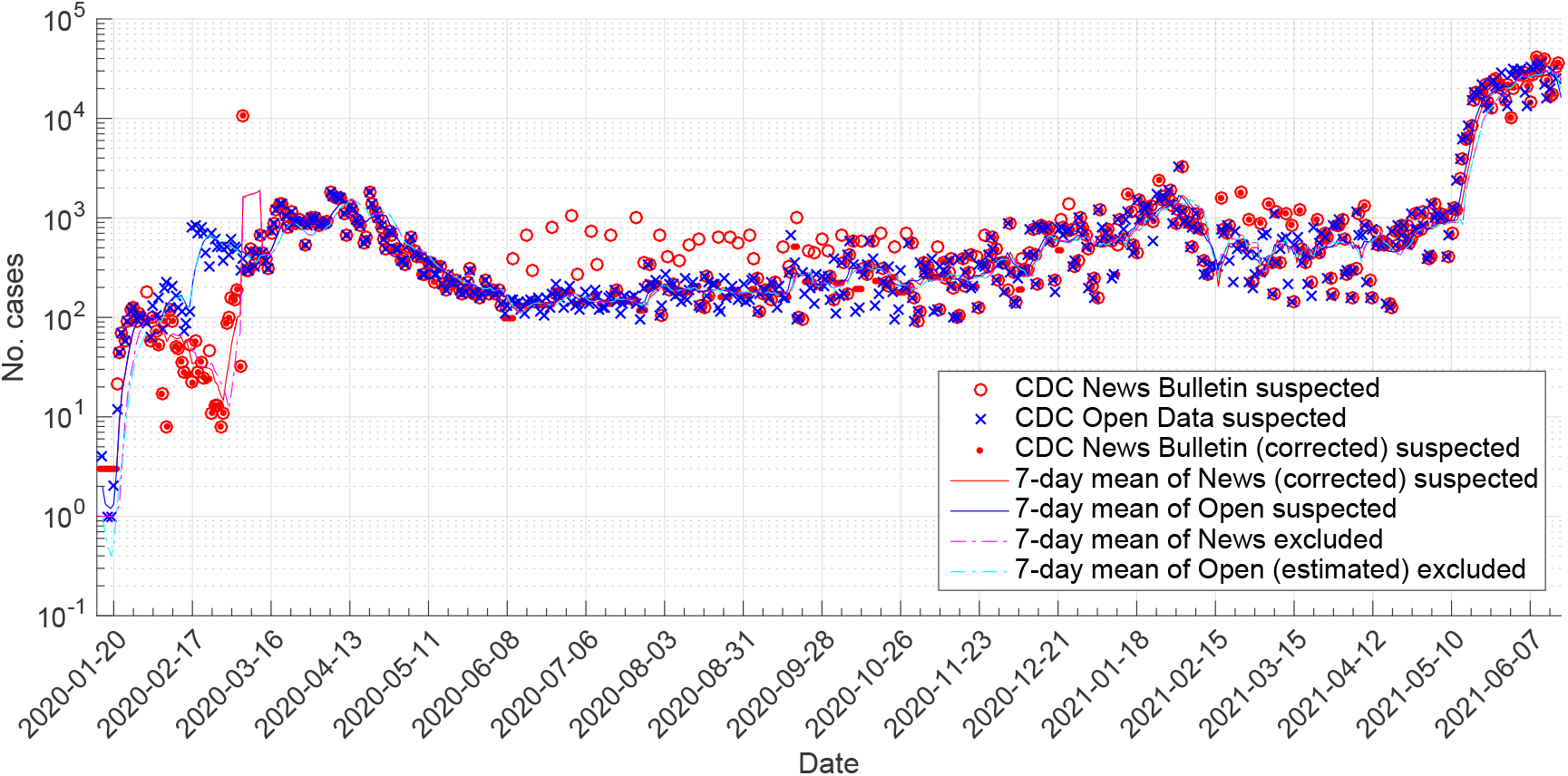
Comparison of the change in number of suspected between CDC News Bulletins (red circles), the total daily number of suspected in the CDC Open Data (blue x), and the change in number of suspected between CDC News Bulletins corrected for the number of days in between (red dot), and the 7-day moving mean of the later two (blue and red lines), and the 7-day moving mean of the change in number of excluded between CDC News Bulletins corrected for the number of days in between (magenta line), and the 7-day moving mean of our estimate of the daily number of excluded based on the total daily number of suspected in the CDC Open Data (cyan line).

**Figure 4:**
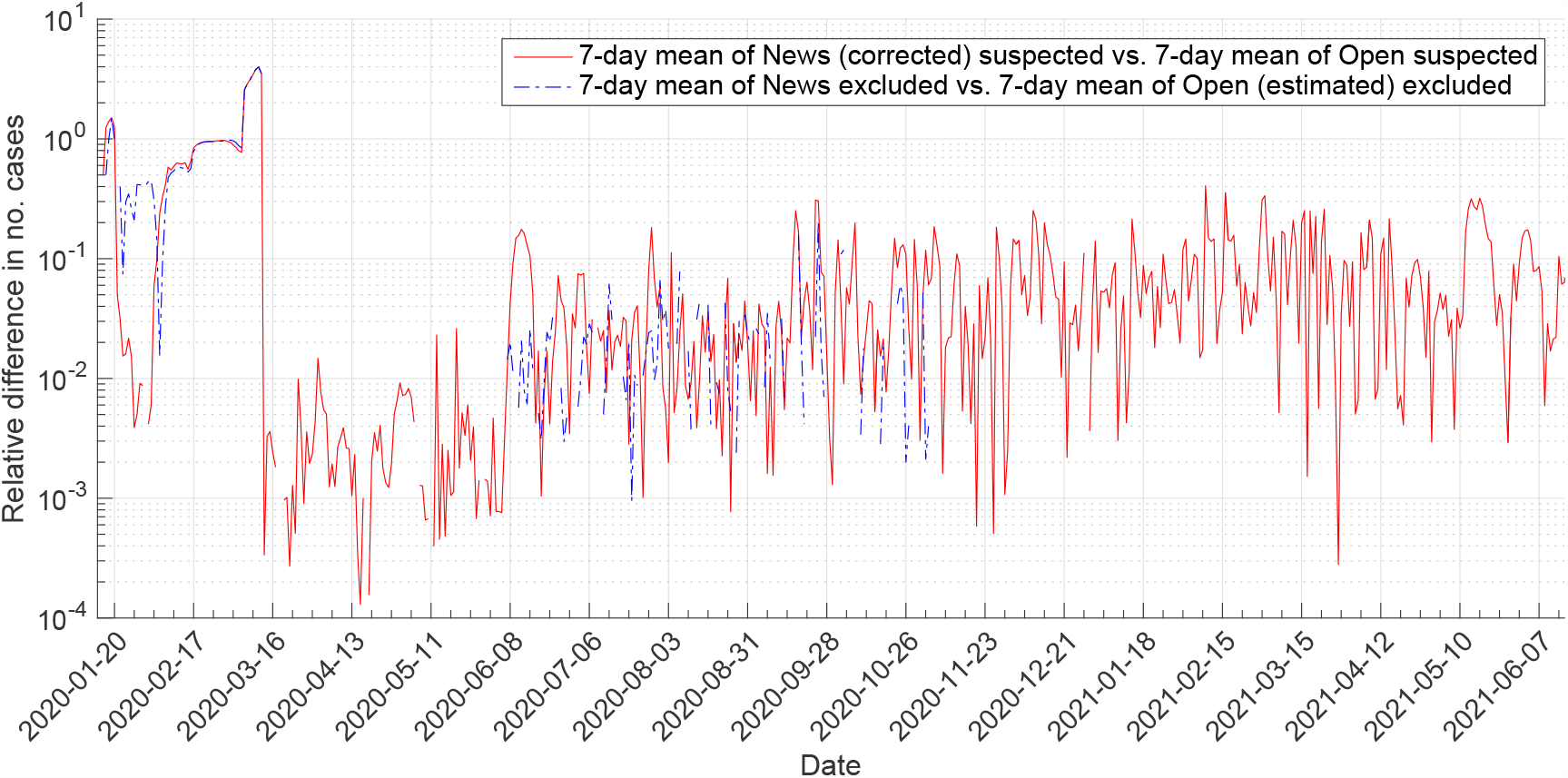
Relative difference in the 7-day moving mean of the change in number of suspected between CDC News Bulletins corrected for the number of days in between and the 7-day moving mean of the total daily number of suspected in the CDC Open Data (red line), and the 7-day moving mean of the change in number of excluded between CDC News Bulletins corrected for the number of days in between and the 7-day moving mean of our estimate of the daily number of excluded based on the total daily number of suspected in the CDC Open Data (magenta line).

Since the number of excluded cases only was available from in the CDC News Bulletins and contain a similar change in the way fo counting on 2020-03-06, we assumed that the ratio of excluded to suspected was correct and used the ratio to estimate the number of daily excluded based on the total number of daily suspected from the CDC Open Data dataset. To ensure that the cumulative sum of daily excluded matched the reported values in the News Bulletins after March 6th, we proportionally to the daily suspected added or removed excluded cases from the days between two reports iteratively from the later report day to the former to avoid errors from rounding to whole integers later. This resulted in an estimate of the daily total number of excluded cases that on each day with a CDC News Bulletin report match the reported number exactly. The result of our preprocessing is visualised in Figure 3, where the number of suspected cases from the corrected News Bulletin and Open Data closely match after 2020-03-06. Note that a red circle without a dot in it indicate that more than one day had passed between the consecutive News Bulletins and the change is therefore larger than expected from one day to the next. We have verified that the cumulative number of excluded always remain smaller than the cumulative number of suspected.

Since our analysis if dependent on the 7-day moving mean of suspected and excluded we also evaluated the relative difference in the 7-day moving mean daily values obtained from the CDC News Bulletin and Open Data, relative to the later. The relative difference in number of suspected remains below 41% every day after 2020-03-06, below 10% 81% of the days, and below 1% 33% of the days. The large relative differences occur on separated days with small relative differences in between and are caused by differences in on which day extreme numbers of suspected are reported, see Figure 3. This is to be expected, since the Open Data most probably contains all suspected cases on each day, while the News Bulletins contain the cases added since the last bulletin. Thus these differences may shift values by one day, but no more. We cannot tell which of these two data sources that is more reliable, but considering that the Open Data at least is corrected for the change in the way of calculating the number of suspected we decided to use it and our estimate of the total daily number of excluded based on it. This does not affect the ratio of excluded suspected cases, but it affects the ratio of local & unknown confirmed to suspected cases by up to 41% on individual days, which in practise means that specific values may be shifted in time by one day. Since our forecast of when the third outbreak will end is based on 19 time points any effects of this difference is well within the estimated confidence intervals.

## Data used

The data we used after cleaning and preprocessing is included in Table 1-4.

**Table 1:** The data used in this study.

| Date | Susp. | Excl. | Conf. | Date | Susp. | Excl. | Conf. | Date | Susp. | Excl. | Conf. |
| --- | --- | --- | --- | --- | --- | --- | --- | --- | --- | --- | --- |
| 2020-01-15 | 0 | 0 | 0 | 2020-03-03 | 506 | 466 | 0 | 2020-04-20 | 1835 | 931 | 0 |
| 2020-01-16 | 4 | 2 | 0 | 2020-03-04 | 485 | 454 | 1 | 2020-04-21 | 1371 | 1526 | 0 |
| 2020-01-17 | 0 | 0 | 0 | 2020-03-05 | 297 | 283 | 1 | 2020-04-22 | 1154 | 1469 | 0 |
| 2020-01-18 | 1 | 0 | 0 | 2020-03-06 | 381 | 393 | 0 | 2020-04-23 | 1014 | 1163 | 0 |
| 2020-01-19 | 1 | 0 | 0 | 2020-03-07 | 307 | 382 | 0 | 2020-04-24 | 827 | 1132 | 0 |
| 2020-01-20 | 2 | 1 | 0 | 2020-03-08 | 298 | 313 | 0 | 2020-04-25 | 619 | 1091 | 0 |
| 2020-01-21 | 12 | 5 | 0 | 2020-03-09 | 476 | 359 | 1 | 2020-04-26 | 495 | 738 | 0 |
| 2020-01-22 | 44 | 7 | 0 | 2020-03-10 | 332 | 376 | 0 | 2020-04-27 | 740 | 588 | 0 |
| 2020-01-23 | 70 | 14 | 0 | 2020-03-11 | 445 | 395 | 0 | 2020-04-28 | 694 | 792 | 0 |
| 2020-01-24 | 57 | 20 | 0 | 2020-03-12 | 677 | 685 | 1 | 2020-04-29 | 484 | 705 | 0 |
| 2020-01-25 | 91 | 42 | 0 | 2020-03-13 | 439 | 454 | 0 | 2020-04-30 | 495 | 513 | 0 |
| 2020-01-26 | 117 | 58 | 0 | 2020-03-14 | 379 | 405 | 0 | 2020-05-01 | 374 | 477 | 0 |
| 2020-01-27 | 123 | 64 | 0 | 2020-03-15 | 306 | 341 | 0 | 2020-05-02 | 380 | 436 | 0 |
| 2020-01-28 | 95 | 62 | 1 | 2020-03-16 | 730 | 388 | 0 | 2020-05-03 | 342 | 365 | 0 |
| 2020-01-29 | 106 | 74 | 1 | 2020-03-17 | 873 | 600 | 2 | 2020-05-04 | 528 | 442 | 0 |
| 2020-01-30 | 100 | 74 | 0 | 2020-03-18 | 1197 | 786 | 1 | 2020-05-05 | 632 | 660 | 0 |
| 2020-01-31 | 93 | 82 | 0 | 2020-03-19 | 1363 | 1095 | 3 | 2020-05-06 | 451 | 507 | 0 |
| 2020-02-01 | 87 | 76 | 0 | 2020-03-20 | 1371 | 1236 | 0 | 2020-05-07 | 417 | 457 | 0 |
| 2020-02-02 | 64 | 57 | 0 | 2020-03-21 | 1047 | 880 | 3 | 2020-05-08 | 402 | 408 | 0 |
| 2020-02-03 | 131 | 116 | 0 | 2020-03-22 | 811 | 823 | 1 | 2020-05-09 | 269 | 386 | 0 |
| 2020-02-04 | 103 | 91 | 0 | 2020-03-23 | 1139 | 734 | 0 | 2020-05-10 | 267 | 293 | 0 |
| 2020-02-05 | 157 | 147 | 0 | 2020-03-24 | 924 | 697 | 1 | 2020-05-11 | 362 | 307 | 0 |
| 2020-02-06 | 77 | 73 | 0 | 2020-03-25 | 938 | 603 | 2 | 2020-05-12 | 349 | 343 | 0 |
| 2020-02-07 | 199 | 191 | 0 | 2020-03-26 | 929 | 802 | 0 | 2020-05-13 | 277 | 317 | 0 |
| 2020-02-08 | 225 | 221 | 0 | 2020-03-27 | 864 | 881 | 2 | 2020-05-14 | 274 | 285 | 0 |
| 2020-02-09 | 128 | 121 | 0 | 2020-03-28 | 527 | 592 | 1 | 2020-05-15 | 323 | 306 | 0 |
| 2020-02-10 | 205 | 195 | 0 | 2020-03-29 | 835 | 456 | 1 | 2020-05-16 | 220 | 283 | 0 |
| 2020-02-11 | 168 | 162 | 0 | 2020-03-30 | 1023 | 679 | 2 | 2020-05-17 | 185 | 214 | 0 |
| 2020-02-12 | 128 | 127 | 0 | 2020-03-31 | 924 | 708 | 0 | 2020-05-18 | 269 | 235 | 0 |
| 2020-02-13 | 115 | 117 | 0 | 2020-04-01 | 1009 | 990 | 2 | 2020-05-19 | 223 | 236 | 0 |
| 2020-02-14 | 72 | 73 | 0 | 2020-04-02 | 822 | 775 | 2 | 2020-05-20 | 226 | 253 | 0 |
| 2020-02-15 | 89 | 91 | 1 | 2020-04-03 | 880 | 854 | 1 | 2020-05-21 | 236 | 247 | 0 |
| 2020-02-16 | 112 | 115 | 1 | 2020-04-04 | 864 | 894 | 0 | 2020-05-22 | 187 | 221 | 0 |
| 2020-02-17 | 813 | 827 | 2 | 2020-04-05 | 912 | 850 | 1 | 2020-05-23 | 170 | 200 | 0 |
| 2020-02-18 | 832 | 841 | 1 | 2020-04-06 | 1814 | 894 | 0 | 2020-05-24 | 183 | 183 | 0 |
| 2020-02-19 | 696 | 706 | 1 | 2020-04-07 | 1685 | 1507 | 1 | 2020-05-25 | 222 | 198 | 0 |
| 2020-02-20 | 796 | 808 | 2 | 2020-04-08 | 1628 | 1725 | 1 | 2020-05-26 | 299 | 204 | 0 |
| 2020-02-21 | 714 | 728 | 0 | 2020-04-09 | 1576 | 1526 | 0 | 2020-05-27 | 183 | 286 | 0 |
| 2020-02-22 | 444 | 459 | 2 | 2020-04-10 | 1564 | 1613 | 0 | 2020-05-28 | 170 | 174 | 0 |
| 2020-02-23 | 328 | 340 | 2 | 2020-04-11 | 1091 | 1288 | 1 | 2020-05-29 | 156 | 169 | 0 |
| 2020-02-24 | 698 | 725 | 1 | 2020-04-12 | 664 | 1026 | 0 | 2020-05-30 | 174 | 168 | 0 |
| 2020-02-25 | 609 | 632 | 1 | 2020-04-13 | 1340 | 2183 | 0 | 2020-05-31 | 237 | 160 | 0 |
| 2020-02-26 | 521 | 541 | 0 | 2020-04-14 | 1204 | 1439 | 0 | 2020-06-01 | 186 | 197 | 0 |
| 2020-02-27 | 520 | 541 | 1 | 2020-04-15 | 952 | 1078 | 0 | 2020-06-02 | 178 | 245 | 0 |
| 2020-02-28 | 371 | 386 | 4 | 2020-04-16 | 903 | 1273 | 0 | 2020-06-03 | 188 | 185 | 0 |
| 2020-02-29 | 511 | 513 | 0 | 2020-04-17 | 838 | 1046 | 0 | 2020-06-04 | 170 | 175 | 0 |
| 2020-03-01 | 430 | 415 | 1 | 2020-04-18 | 557 | 893 | 0 | 2020-06-05 | 184 | 201 | 0 |
| 2020-03-02 | 600 | 559 | 1 | 2020-04-19 | 624 | 600 | 0 | 2020-06-06 | 138 | 177 | 0 |

**Table 2:** The data used in this study (continued).

| Date | Susp. | Excl. | Conf. | Date | Susp. | Excl. | Conf. | Date | Susp. | Excl. | Conf. |
| --- | --- | --- | --- | --- | --- | --- | --- | --- | --- | --- | --- |
| 2020-06-07 | 108 | 71 | 0 | 2020-07-25 | 95 | 85 | 0 | 2020-09-11 | 186 | 205 | 0 |
| 2020-06-08 | 129 | 84 | 0 | 2020-07-26 | 123 | 112 | 0 | 2020-09-12 | 126 | 123 | 0 |
| 2020-06-09 | 152 | 101 | 0 | 2020-07-27 | 201 | 182 | 0 | 2020-09-13 | 210 | 205 | 0 |
| 2020-06-10 | 152 | 101 | 0 | 2020-07-28 | 341 | 143 | 0 | 2020-09-14 | 154 | 152 | 0 |
| 2020-06-11 | 135 | 141 | 0 | 2020-07-29 | 220 | 320 | 0 | 2020-09-15 | 160 | 137 | 0 |
| 2020-06-12 | 142 | 148 | 0 | 2020-07-30 | 203 | 232 | 0 | 2020-09-16 | 282 | 244 | 0 |
| 2020-06-13 | 118 | 125 | 0 | 2020-07-31 | 235 | 271 | 0 | 2020-09-17 | 680 | 177 | 0 |
| 2020-06-14 | 111 | 117 | 0 | 2020-08-01 | 104 | 120 | 1 | 2020-09-18 | 351 | 793 | 0 |
| 2020-06-15 | 145 | 152 | 0 | 2020-08-02 | 132 | 196 | 0 | 2020-09-19 | 95 | 216 | 0 |
| 2020-06-16 | 147 | 134 | 0 | 2020-08-03 | 273 | 178 | 0 | 2020-09-20 | 98 | 204 | 0 |
| 2020-06-17 | 146 | 134 | 0 | 2020-08-04 | 192 | 126 | 0 | 2020-09-21 | 283 | 89 | 0 |
| 2020-06-18 | 135 | 164 | 0 | 2020-08-05 | 212 | 215 | 0 | 2020-09-22 | 175 | 174 | 0 |
| 2020-06-19 | 159 | 152 | 0 | 2020-08-06 | 214 | 182 | 0 | 2020-09-23 | 257 | 256 | 0 |
| 2020-06-20 | 114 | 109 | 0 | 2020-08-07 | 164 | 200 | 0 | 2020-09-24 | 198 | 293 | 0 |
| 2020-06-21 | 104 | 99 | 0 | 2020-08-08 | 173 | 212 | 0 | 2020-09-25 | 137 | 203 | 0 |
| 2020-06-22 | 156 | 149 | 0 | 2020-08-09 | 142 | 194 | 0 | 2020-09-26 | 275 | 235 | 0 |
| 2020-06-23 | 126 | 121 | 0 | 2020-08-10 | 247 | 230 | 0 | 2020-09-27 | 204 | 175 | 0 |
| 2020-06-24 | 173 | 166 | 0 | 2020-08-11 | 147 | 137 | 0 | 2020-09-28 | 259 | 222 | 0 |
| 2020-06-25 | 217 | 129 | 0 | 2020-08-12 | 190 | 178 | 0 | 2020-09-29 | 211 | 176 | 0 |
| 2020-06-26 | 176 | 188 | 0 | 2020-08-13 | 229 | 235 | 0 | 2020-09-30 | 260 | 218 | 0 |
| 2020-06-27 | 154 | 164 | 0 | 2020-08-14 | 198 | 204 | 0 | 2020-10-01 | 152 | 176 | 0 |
| 2020-06-28 | 127 | 135 | 0 | 2020-08-15 | 133 | 137 | 0 | 2020-10-02 | 392 | 187 | 0 |
| 2020-06-29 | 184 | 198 | 0 | 2020-08-16 | 124 | 180 | 0 | 2020-10-03 | 109 | 121 | 0 |
| 2020-06-30 | 210 | 225 | 0 | 2020-08-17 | 260 | 116 | 0 | 2020-10-04 | 167 | 187 | 0 |
| 2020-07-01 | 141 | 151 | 0 | 2020-08-18 | 165 | 216 | 0 | 2020-10-05 | 356 | 398 | 0 |
| 2020-07-02 | 129 | 143 | 0 | 2020-08-19 | 222 | 204 | 0 | 2020-10-06 | 170 | 174 | 0 |
| 2020-07-03 | 140 | 157 | 0 | 2020-08-20 | 214 | 236 | 0 | 2020-10-07 | 432 | 242 | 0 |
| 2020-07-04 | 119 | 118 | 0 | 2020-08-21 | 217 | 239 | 0 | 2020-10-08 | 590 | 457 | 0 |
| 2020-07-05 | 151 | 149 | 0 | 2020-08-22 | 128 | 142 | 0 | 2020-10-09 | 339 | 420 | 0 |
| 2020-07-06 | 177 | 175 | 0 | 2020-08-23 | 117 | 111 | 0 | 2020-10-10 | 114 | 184 | 0 |
| 2020-07-07 | 136 | 135 | 0 | 2020-08-24 | 242 | 230 | 0 | 2020-10-11 | 123 | 199 | 0 |
| 2020-07-08 | 141 | 140 | 0 | 2020-08-25 | 155 | 147 | 0 | 2020-10-12 | 305 | 492 | 0 |
| 2020-07-09 | 188 | 144 | 0 | 2020-08-26 | 182 | 173 | 0 | 2020-10-13 | 271 | 316 | 0 |
| 2020-07-10 | 165 | 128 | 0 | 2020-08-27 | 204 | 210 | 0 | 2020-10-14 | 345 | 267 | 0 |
| 2020-07-11 | 118 | 126 | 0 | 2020-08-28 | 177 | 183 | 0 | 2020-10-15 | 589 | 282 | 0 |
| 2020-07-12 | 126 | 133 | 0 | 2020-08-29 | 148 | 153 | 0 | 2020-10-16 | 382 | 365 | 0 |
| 2020-07-13 | 126 | 134 | 0 | 2020-08-30 | 157 | 166 | 0 | 2020-10-17 | 132 | 195 | 0 |
| 2020-07-14 | 155 | 166 | 0 | 2020-08-31 | 215 | 229 | 0 | 2020-10-18 | 176 | 262 | 0 |
| 2020-07-15 | 164 | 175 | 0 | 2020-09-01 | 144 | 153 | 0 | 2020-10-19 | 314 | 467 | 0 |
| 2020-07-16 | 191 | 152 | 0 | 2020-09-02 | 152 | 162 | 0 | 2020-10-20 | 223 | 209 | 0 |
| 2020-07-17 | 145 | 196 | 0 | 2020-09-03 | 232 | 181 | 0 | 2020-10-21 | 215 | 180 | 0 |
| 2020-07-18 | 111 | 107 | 0 | 2020-09-04 | 242 | 189 | 0 | 2020-10-22 | 329 | 214 | 0 |
| 2020-07-19 | 135 | 129 | 0 | 2020-09-05 | 112 | 186 | 0 | 2020-10-23 | 177 | 353 | 0 |
| 2020-07-20 | 161 | 155 | 0 | 2020-09-06 | 173 | 204 | 0 | 2020-10-24 | 97 | 195 | 0 |
| 2020-07-21 | 143 | 137 | 0 | 2020-09-07 | 188 | 91 | 0 | 2020-10-25 | 118 | 103 | 0 |
| 2020-07-22 | 156 | 151 | 0 | 2020-09-08 | 187 | 221 | 0 | 2020-10-26 | 293 | 256 | 0 |
| 2020-07-23 | 163 | 158 | 0 | 2020-09-09 | 155 | 185 | 0 | 2020-10-27 | 206 | 181 | 0 |
| 2020-07-24 | 135 | 130 | 0 | 2020-09-10 | 224 | 146 | 0 | 2020-10-28 | 198 | 174 | 0 |

**Table 3:** The data used in this study (continued).

| Date | Susp. | Excl. | Conf. | Date | Susp. | Excl. | Conf. | Date | Susp. | Excl. | Conf. |
| --- | --- | --- | --- | --- | --- | --- | --- | --- | --- | --- | --- |
| 2020-10-29 | 569 | 192 | 0 | 2020-12-16 | 757 | 844 | 0 | 2021-02-02 | 3243 | 1000 | 0 |
| 2020-10-30 | 156 | 244 | 0 | 2020-12-17 | 790 | 715 | 0 | 2021-02-03 | 1167 | 2029 | 0 |
| 2020-10-31 | 93 | 208 | 0 | 2020-12-18 | 577 | 573 | 0 | 2021-02-04 | 927 | 1082 | 0 |
| 2020-11-01 | 116 | 191 | 0 | 2020-12-19 | 240 | 502 | 0 | 2021-02-05 | 877 | 2127 | 1 |
| 2020-11-02 | 352 | 237 | 0 | 2020-12-20 | 183 | 559 | 0 | 2021-02-06 | 517 | 1918 | 0 |
| 2020-11-03 | 288 | 262 | 0 | 2020-12-21 | 760 | 432 | 0 | 2021-02-07 | 793 | 825 | 0 |
| 2020-11-04 | 325 | 259 | 0 | 2020-12-22 | 800 | 455 | 1 | 2021-02-08 | 1107 | 457 | 0 |
| 2020-11-05 | 272 | 262 | 0 | 2020-12-23 | 670 | 851 | 0 | 2021-02-09 | 784 | 844 | 1 |
| 2020-11-06 | 394 | 230 | 0 | 2020-12-24 | 728 | 560 | 0 | 2021-02-10 | 379 | 729 | 0 |
| 2020-11-07 | 114 | 209 | 0 | 2020-12-25 | 759 | 586 | 0 | 2021-02-11 | 275 | 639 | 0 |
| 2020-11-08 | 118 | 217 | 0 | 2020-12-26 | 324 | 678 | 0 | 2021-02-12 | 276 | 369 | 0 |
| 2020-11-09 | 320 | 180 | 0 | 2020-12-27 | 365 | 624 | 0 | 2021-02-13 | 324 | 0 | 0 |
| 2020-11-10 | 292 | 262 | 0 | 2020-12-28 | 1017 | 356 | 0 | 2021-02-14 | 334 | 0 | 0 |
| 2020-11-11 | 287 | 388 | 0 | 2020-12-29 | 810 | 770 | 0 | 2021-02-15 | 325 | 0 | 0 |
| 2020-11-12 | 380 | 363 | 0 | 2020-12-30 | 674 | 778 | 0 | 2021-02-16 | 320 | 0 | 0 |
| 2020-11-13 | 204 | 296 | 0 | 2020-12-31 | 507 | 745 | 0 | 2021-02-17 | 759 | 2001 | 0 |
| 2020-11-14 | 98 | 201 | 0 | 2021-01-01 | 200 | 621 | 0 | 2021-02-18 | 698 | 632 | 0 |
| 2020-11-15 | 102 | 149 | 0 | 2021-01-02 | 251 | 402 | 0 | 2021-02-19 | 839 | 821 | 0 |
| 2020-11-16 | 382 | 182 | 0 | 2021-01-03 | 159 | 302 | 0 | 2021-02-20 | 371 | 839 | 0 |
| 2020-11-17 | 215 | 238 | 0 | 2021-01-04 | 1185 | 180 | 0 | 2021-02-21 | 231 | 0 | 0 |
| 2020-11-18 | 287 | 286 | 0 | 2021-01-05 | 597 | 654 | 0 | 2021-02-22 | 736 | 0 | 0 |
| 2020-11-19 | 429 | 277 | 0 | 2021-01-06 | 807 | 987 | 0 | 2021-02-23 | 487 | 0 | 0 |
| 2020-11-20 | 199 | 265 | 0 | 2021-01-07 | 699 | 603 | 0 | 2021-02-24 | 546 | 2445 | 0 |
| 2020-11-21 | 210 | 325 | 0 | 2021-01-08 | 597 | 728 | 0 | 2021-02-25 | 544 | 525 | 0 |
| 2020-11-22 | 126 | 195 | 0 | 2021-01-09 | 249 | 539 | 0 | 2021-02-26 | 418 | 0 | 0 |
| 2020-11-23 | 341 | 200 | 0 | 2021-01-10 | 276 | 176 | 0 | 2021-02-27 | 221 | 669 | 0 |
| 2020-11-24 | 342 | 240 | 0 | 2021-01-11 | 985 | 628 | 0 | 2021-02-28 | 197 | 307 | 0 |
| 2020-11-25 | 329 | 232 | 0 | 2021-01-12 | 612 | 697 | 2 | 2021-03-01 | 276 | 0 | 0 |
| 2020-11-26 | 425 | 320 | 0 | 2021-01-13 | 758 | 862 | 0 | 2021-03-02 | 434 | 0 | 0 |
| 2020-11-27 | 638 | 392 | 0 | 2021-01-14 | 967 | 0 | 0 | 2021-03-03 | 288 | 789 | 0 |
| 2020-11-28 | 479 | 359 | 0 | 2021-01-15 | 1174 | 1446 | 0 | 2021-03-04 | 675 | 278 | 0 |
| 2020-11-29 | 241 | 693 | 0 | 2021-01-16 | 543 | 836 | 0 | 2021-03-05 | 706 | 0 | 0 |
| 2020-11-30 | 337 | 377 | 0 | 2021-01-17 | 447 | 854 | 2 | 2021-03-06 | 349 | 1225 | 0 |
| 2020-12-01 | 363 | 280 | 0 | 2021-01-18 | 1518 | 610 | 1 | 2021-03-07 | 176 | 771 | 0 |
| 2020-12-02 | 184 | 262 | 0 | 2021-01-19 | 951 | 657 | 4 | 2021-03-08 | 1093 | 233 | 0 |
| 2020-12-03 | 898 | 301 | 0 | 2021-01-20 | 1244 | 1178 | 1 | 2021-03-09 | 357 | 551 | 0 |
| 2020-12-04 | 283 | 280 | 0 | 2021-01-21 | 1188 | 1202 | 0 | 2021-03-10 | 490 | 880 | 0 |
| 2020-12-05 | 146 | 605 | 0 | 2021-01-22 | 1324 | 1341 | 2 | 2021-03-11 | 626 | 0 | 0 |
| 2020-12-06 | 137 | 352 | 0 | 2021-01-23 | 1007 | 1081 | 1 | 2021-03-12 | 641 | 799 | 0 |
| 2020-12-07 | 242 | 141 | 0 | 2021-01-24 | 593 | 1293 | 2 | 2021-03-13 | 212 | 0 | 0 |
| 2020-12-08 | 291 | 170 | 0 | 2021-01-25 | 1738 | 0 | 0 | 2021-03-14 | 142 | 1034 | 0 |
| 2020-12-09 | 300 | 211 | 0 | 2021-01-26 | 1516 | 1867 | 0 | 2021-03-15 | 536 | 152 | 0 |
| 2020-12-10 | 434 | 78 | 0 | 2021-01-27 | 1629 | 1639 | 0 | 2021-03-16 | 650 | 0 | 0 |
| 2020-12-11 | 780 | 330 | 0 | 2021-01-28 | 1602 | 1290 | 0 | 2021-03-17 | 485 | 865 | 0 |
| 2020-12-12 | 423 | 540 | 0 | 2021-01-29 | 1878 | 1609 | 0 | 2021-03-18 | 360 | 479 | 0 |
| 2020-12-13 | 216 | 1065 | 0 | 2021-01-30 | 1359 | 1362 | 4 | 2021-03-19 | 432 | 713 | 0 |
| 2020-12-14 | 847 | 270 | 0 | 2021-01-31 | 872 | 1277 | 0 | 2021-03-20 | 219 | 424 | 0 |
| 2020-12-15 | 801 | 594 | 0 | 2021-02-01 | 1210 | 1220 | 0 | 2021-03-21 | 163 | 391 | 0 |

**Table 4:** The data used in this study (continued).

| Date | Susp. | Excl. | Conf. | Date | Susp. | Excl. | Conf | Date | Susp. | Excl. | Conf |
| --- | --- | --- | --- | --- | --- | --- | --- | --- | --- | --- | --- |
| 2021-03-22 | 803 | 0 | 0 | 2021-04-20 | 526 | 413 | 2 | 2021-05-19 | 17528 | 8746 | 265 |
| 2021-03-23 | 437 | 771 | 0 | 2021-04-21 | 837 | 577 | 0 | 2021-05-20 | 17892 | 12403 | 283 |
| 2021-03-24 | 569 | 706 | 0 | 2021-04-22 | 670 | 520 | 0 | 2021-05-21 | 21653 | 15358 | 311 |
| 2021-03-25 | 716 | 556 | 0 | 2021-04-23 | 832 | 920 | 2 | 2021-05-22 | 14450 | 17597 | 721 |
| 2021-03-26 | 628 | 763 | 0 | 2021-04-24 | 543 | 656 | 1 | 2021-05-23 | 12865 | 14176 | 457 |
| 2021-03-27 | 252 | 637 | 0 | 2021-04-25 | 614 | 479 | 2 | 2021-05-24 | 22926 | 12381 | 590 |
| 2021-03-28 | 164 | 405 | 0 | 2021-04-26 | 1099 | 488 | 2 | 2021-05-25 | 24383 | 15368 | 537 |
| 2021-03-29 | 850 | 212 | 0 | 2021-04-27 | 752 | 1089 | 1 | 2021-05-26 | 20871 | 18701 | 633 |
| 2021-03-30 | 834 | 604 | 0 | 2021-04-28 | 764 | 1018 | 3 | 2021-05-27 | 19811 | 22596 | 666 |
| 2021-03-31 | 964 | 807 | 0 | 2021-04-29 | 1144 | 685 | 3 | 2021-05-28 | 28993 | 15799 | 552 |
| 2021-04-01 | 604 | 623 | 0 | 2021-04-30 | 854 | 1056 | 3 | 2021-05-29 | 16141 | 29381 | 484 |
| 2021-04-02 | 275 | 693 | 0 | 2021-05-01 | 383 | 766 | 1 | 2021-05-30 | 13535 | 27136 | 354 |
| 2021-04-03 | 290 | 412 | 0 | 2021-05-02 | 418 | 625 | 4 | 2021-05-31 | 27502 | 11967 | 347 |
| 2021-04-04 | 158 | 362 | 0 | 2021-05-03 | 1393 | 435 | 2 | 2021-06-01 | 30905 | 22678 | 326 |
| 2021-04-05 | 303 | 171 | 0 | 2021-05-04 | 988 | 993 | 2 | 2021-06-02 | 29598 | 35689 | 548 |
| 2021-04-06 | 1159 | 421 | 0 | 2021-05-05 | 1094 | 1194 | 0 | 2021-06-03 | 27796 | 31670 | 583 |
| 2021-04-07 | 658 | 478 | 0 | 2021-05-06 | 1006 | 1001 | 1 | 2021-06-04 | 31416 | 29217 | 470 |
| 2021-04-08 | 669 | 0 | 0 | 2021-05-07 | 1067 | 939 | 0 | 2021-06-05 | 18505 | 29470 | 510 |
| 2021-04-09 | 846 | 1716 | 0 | 2021-05-08 | 413 | 926 | 2 | 2021-06-06 | 13167 | 20020 | 342 |
| 2021-04-10 | 230 | 658 | 0 | 2021-05-09 | 680 | 698 | 0 | 2021-06-07 | 33272 | 14079 | 190 |
| 2021-04-11 | 159 | 553 | 0 | 2021-05-10 | 1237 | 556 | 4 | 2021-06-08 | 31728 | 26995 | 203 |
| 2021-04-12 | 755 | 344 | 0 | 2021-05-11 | 1218 | 1003 | 7 | 2021-06-09 | 33580 | 41180 | 273 |
| 2021-04-13 | 538 | 638 | 0 | 2021-05-12 | 2432 | 1136 | 17 | 2021-06-10 | 35025 | 29801 | 251 |
| 2021-04-14 | 598 | 721 | 0 | 2021-05-13 | 3893 | 1647 | 13 | 2021-06-11 | 36148 | 34517 | 277 |
| 2021-04-15 | 526 | 570 | 0 | 2021-05-14 | 6177 | 0 | 29 | 2021-06-12 | 22323 | 40622 | 245 |
| 2021-04-16 | 548 | 507 | 0 | 2021-05-15 | 6497 | 7736 | 180 | 2021-06-13 | 15821 | 24841 | 174 |
| 2021-04-17 | 139 | 511 | 0 | 2021-05-16 | 8425 | 4755 | 206 | 2021-06-14 | 19907 | 16906 | 185 |
| 2021-04-18 | 128 | 319 | 0 | 2021-05-17 | 15447 | 6322 | 333 | 2021-06-15 | 30647 | 17792 | 132 |
| 2021-04-19 | 719 | 133 | 0 | 2021-05-18 | 18087 | 8306 | 238 | 2021-06-16 | 25689 | 33366 | 165 |

## Bibliography

[1] Hsiao YH, Hetherington W. Virus Outbreak: Masks required for train passengers. Taipei, Taiwan: The Taipei Times; 2020. Available from: https://www.taipeitimes.com/News/ taiwan/archives/2020/04/01/2003733786.

[2] Taiwan News. 5 Taiwanese attend Mayday concert despite self-monitoring for COVID. Taipei, Taiwan: Taiwan News; 2021. Available from: https://www.taiwannews.com.tw/ en/news/4091789.

[3] Hsu CY, Wang JT, Huang KC, Fan ACH, Yeh YP, Chen SLS. Household transmission but without the community-acquired outbreak of COVID-19 in Taiwan. Journal of the Formosan Medical Association = Taiwan yi zhi. 2021 may;.

[4] Davies NG, Abbott S, Barnard RC, Jarvis CI, Kucharski AJ, Munday JD, et al. Estimated transmissibility and impact of SARS-CoV-2 lineage B. 1.1. 7 in England. Science. 2021;372(6538).

[5] Wang CJ, Ng CY, Brook RH. Response to COVID-19 in Taiwan. JAMA. 2020 apr;323(14):1341. Available from: https://jamanetwork.com/journals/jama/ fullarticle/2762689.

[6] Zennie M. How a False Sense of Security, and a Little Secret Tea, Broke Down Taiwan’s COVID-19 Defenses. Taipei, Taiwan: Time; 2021. Available from: https://time.com/ 6050316/taiwan-covid-19-outbreak-tea/.

[7] Yeh J. CORONAVIRUS/Worried by rise in COVID-19 cases, Taiwan bans large scale gatherings. Taipei, Taiwan: Central News Agency; 2021. Available from: https://focustaiwan.tw/society/202105110007.

[8] Lee IC. Virus Outbreak: CECC urges vigilance as curbs eased. Taipei, Taiwan: The Taipei Times; 2020. Available from: https://www.taipeitimes.com/News/front/archives/ 2020/06/08/2003737829.

[9] Chiang Hc, Chen Cl, Yeh J. CORONAVIRUS/Level 3 COVID-19 alert introduced across Taiwan (Update). Taipei, Taiwan: Central News Agency; 2021. Available from: https://focustaiwan.tw/society/202105190017.

[10] Everington K. Taiwan’s 4 epidemic warning levels. Taipei, Taiwan: Taiwan News; 2021. Available from: https://www.taiwannews.com.tw/en/news/4203460.

[11] Lee JK, Bullen C, Ben Amor Y, Bush SR, Colombo F, Gaviria A, et al. Institutional and behaviour-change interventions to support COVID-19 public health measures: a review by the Lancet Commission Task Force on public health measures to suppress the pandemic. International health. 2021 may;.

[12] Hu C. How to use SMS contact tracing system, powered by Audrey Tang, to enter every store; 2021. Last visited on 2021-06-15. Available from: https://meet-global.bnext.com.tw/articles/view/47383.

[13] Johns Hopkins University of Medicine CRC. Understanding vaccination progress; 2021. Last visited on 2021-06-15. Available from: https://coronavirus.jhu.edu/vaccines/ international.

[14] Staff T. Back to normal: Israel lifts nearly all COVID restraints as virus fades away. Jerusalem, Israel: The Times of Israel; 2021. Available from: https://www.timesofisrael.com/back-to-normal-israel-lifts-nearly-all-covid-restraints-as-virus-fades-away/.

[15] Moghadas SM, Sah P, Vilches TN, Galvani AP. Can the USA return to pre-COVID-19 normal by July 4? The Lancet Infectious Diseases. 2021 jun;Available from: https://linkinghub.elsevier.com/retrieve/pii/S1473309921003248.

[16] Taiwan Centers of Disease Control. Taiwan Centers of Disease Control - Datasets; 2021. Last visited on 2021-06-17. Available from: https://data.cdc.gov.tw/en/dataset.

[17] Taiwan Centers of Disease Control. Taiwan Centers of Disease Control - News Bulletin; 2021. Last visited on 2021-06-17. Available from: https://www.cdc.gov.tw/Bulletin/ List/MmgtpeidAR5Ooai4-fgHzQ.

[18] Chen YC, Yang HP, Li HC, Huang PY, Chen CL, Chiu CH. Features and transmission dynamics of SARS-CoV-2 superspreading events in Taiwan: Implications for effective and sustainable community-centered control. Pediatrics and neonatology. 2021 may;.

